# Long Term Accuracy of SARS-CoV-2 Interferon-γ Release Assay and its Application in Household Investigation

**DOI:** 10.1101/2021.09.20.21263527

**Authors:** Kanagavel Murugesan, Prasanna Jagannathan, Jonathan Altamirano, Yvonne A Maldonado, Hector F. Bonilla, Karen B. Jacobson, Julie Parsonnet, Jason R. Andrews, Run-Zhang Shi, Scott Boyd, Benjamin A. Pinsky, Upinder Singh, Niaz Banaei

**Author notes:** **Corresponding Author:** Niaz Banaei MD, 3375 Hillview Ave, Rm. 1602, Palo Alto, Ca 94304. **Alternative corresponding author:** Kanagavel Murugesan PhD, 3375 Hillview Ave, Palo Alto, Ca 94304.

## Abstract

**Background:** An immunodiagnostic assay that sensitively detects a cell-mediated immune response to SARS-CoV-2 is needed for epidemiological investigation and for clinical assessment of T cell-mediated immune response to vaccines, particularly in the context of emerging variants that might escape antibody responses.

**Methods:** The performance of a whole blood interferon-gamma (IFN-γ) release assay (IGRA) for the detection of SARS-CoV-2 antigen-specific CD4 and CD8 T cells was evaluated in COVID-19 convalescents tested serially up to 10 months post-infection and in healthy blood donors. SARS-CoV-2 IGRA was applied in contacts of households with index cases. Freshly collected blood in the lithium heparin tube was left unstimulated, stimulated with a SARS-CoV-2 peptide pool, and stimulated with mitogen.

**Results:** The overall sensitivity and specificity of IGRA were 84.5% (153/181; 95% confidence interval [CI] 79.0-89.0) and 86.6% (123/142; 95% CI;80.0-91.2), respectively. The sensitivity declined from 100% (16/16; 95% CI 80.6-100) at 0.5-month post-infection to 79.5% (31/39; 95% CI 64.4-89.2) at 10 months post-infection (P<0.01). The IFN-γ response remained relatively robust at 10 months post-infection (3.8 vs. 1.3 IU/mL, respectively). In 14 households, IGRA showed a positivity rate of 100% (12/12) and 65.2% (15/23), and IgG of 50.0% (6/12) and 43.5% (10/23) in index cases and contacts, respectively, exhibiting a difference of +50% (95% CI +25.4-+74.6) and +21.7% (95% CI, +9.23-+42.3), respectively. Either IGRA or IgG was positive in 100% (12/12) of index cases and 73.9% (17/23) of contacts.

**Conclusions:** The SARS-CoV-2 IGRA is a useful clinical diagnostic tool for assessing cell-mediated immune response to SARS-CoV-2.

**Key points:** SARS-CoV-2 immunodiagnostics are needed to identify infected individuals in order to understand the transmission dynamics of emerging variants and to assess vaccine response. Interferon-gamma release assay maintains sensitivity 10 months post-infection in convalescents and detects more household contacts than IgG.

## Introduction

The emergence of severe acute respiratory syndrome coronavirus 2 (SARS-CoV-2) has demanded the development of sensitive laboratory diagnostics to detect active and remote infections to control the pandemic. In the arena of immunodiagnostics, serologic assays that probe antibody responses to SARS-CoV-2 are far more utilized than assays that measure T cell responses. However, serological tests for anti-SARS-CoV-2 antibodies may not accurately predict the magnitude and durability of T cell-mediated immune response to SARS-CoV-2, [1], particularly in immunocompromised patients with impaired B cell function [2, 3]. Recent studies have shown that T cell responses are more sensitive markers of past SARS-CoV-2 infection compared with antibody responses and postulated to represent a correlate of protective immunity [4, 5]. Furthermore, T cell responses are more robust than antibody responses in convalescents with mild or asymptomatic COVID-19 infection [6, 7]. Thus, clinical T cell assays for SARS-CoV-2 are needed to evaluate individuals for the cell-mediated immune response against SARS-CoV-2 as evidence for past infection and immune response to vaccination [8].

Interferon gamma release assay (IGRA) is an in vitro blood diagnostic used clinically to measure IFN-γ released by antigen-specific T cells after stimulation with pathogen-specific peptides. IGRA is best known for its role in diagnosing latent *Mycobacterium tuberculosis* infection [9, 10] in which either purified or whole blood mononuclear cells are stimulated overnight followed by IFN-γ enzyme-linked immunosorbent spot or ELISA, respectively, to measure IFN-γ response from sensitized T cells [11].

Recently, we and another group reported on the accuracy of a laboratory-developed whole blood IGRA for the detection of the SARS-CoV-2 specific T cell responses in convalescents weeks after infection [12, 13]. Since then, our assay has been further optimized with commercial peptide pools and simplified with in-tube stimulation, and offered clinically in a CLIA laboratory at Stanford Health Care. In this study, we report the longitudinal accuracy of SARS-CoV-2 IGRA up to 10 months after infection and evaluated its utility in household contacts exposed to an index case. We show that IGRA response in convalescents is durable and it identifies more infected household contacts compared with antibody testing.

## Methods

### Ethics

The two-parent studies which collected blood from infected and exposed individuals were approved by the Stanford University Institutional Review Board. Informed consent was obtained before blood collection.

### Study design

This was a case-control study to evaluate the longitudinal sensitivity and specificity of SARS-CoV-2 IGRA (Figure 1). Additionally, the utility of SARS-CoV-2 IGRA was assessed in household contacts of COVID-19 index cases. Blood samples from outpatient COVID-19 patients with positive SARS-CoV-2 reverse transcriptase (RT)-PCR enrolled in an IFN-λ vs. placebo therapeutic clinical trial (referred to as cases) and healthy blood donors with no COVID-19 symptoms (referred to as controls) were tested with SARS-CoV-2 IGRA and IgG ELISA. Cases were tested at 0.5, 1, 4, 7 and 10 months post-infection between April 6, 2020 and April 30, 2021. Controls were tested once between May 11, 2020 and November 11, 2021. Based on prior studies, IFN-λ has been shown to not negatively impact adaptive response to SARS-CoV-2 [14]. Household contacts of RT-PCR-positive COVID-19 index cases with mild COVID-19 from 14 households were tested with SARS-CoV-2 IGRA and IgG ELISA days after the index case was diagnosed. Members of each household were tested on the same day.

**Figure 1.**
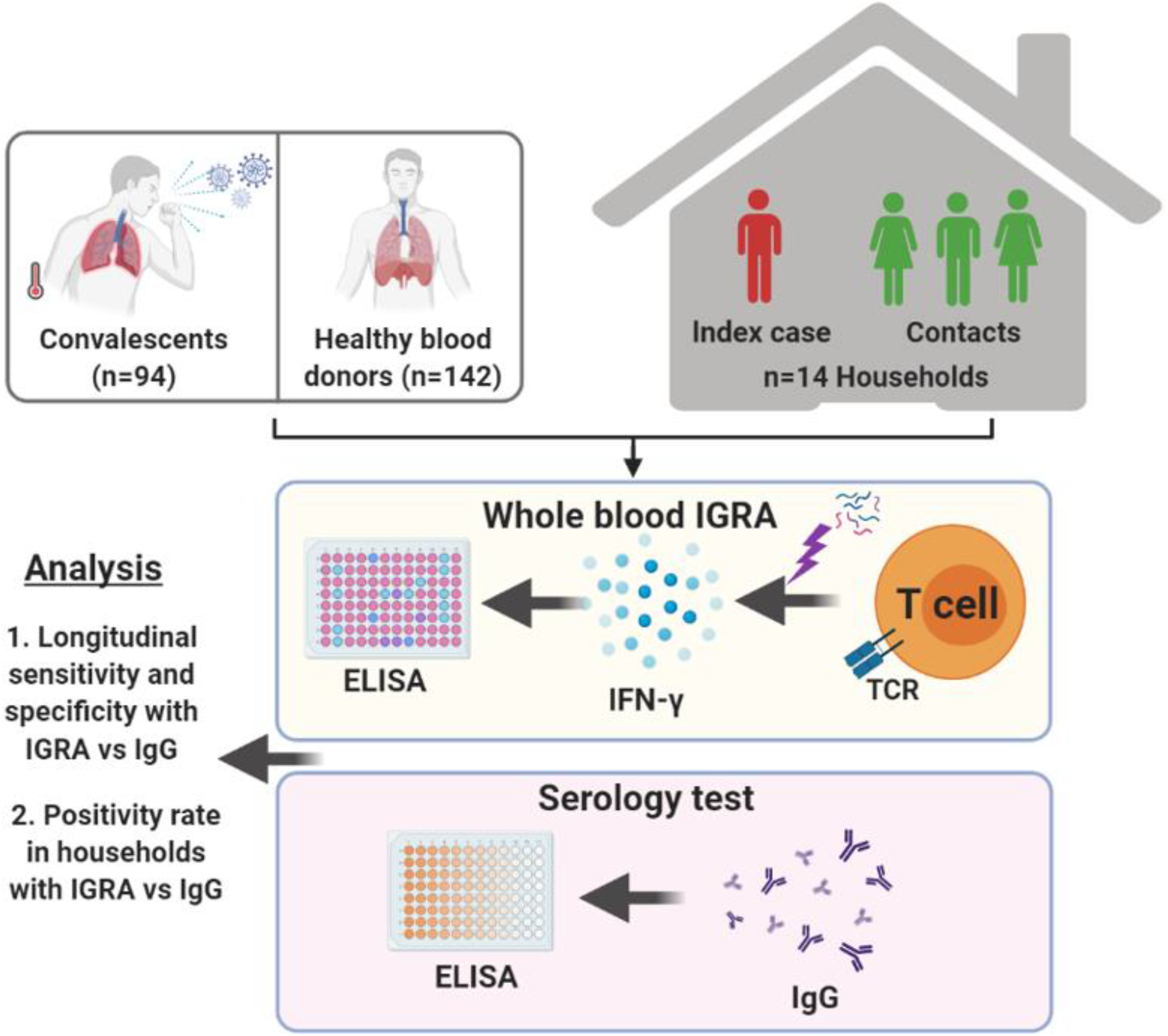
Patient cohorts included in this study to evaluate the performance of whole blood SARS-CoV-2 IGRA.

### Specimens

Whole blood was collected in lithium heparin blood collection tube and transported to the Stanford Health Care Clinical Microbiology Laboratory at room temperature for IGRA testing. Blood from healthy blood donors was purchased from Stanford Blood Center.

### Antigen and Mitogen

The SARS-CoV2 peptide pools consisting of PepTivator SARS-CoV-2 Prot S, S1, N and M were purchased from Miltenyi Biotec (Bergisch Gladbach, Germany). Per the manufacturer, S1 pool consists of 15-mer peptides overlapping by 11-residues covering the S1, while the remaining pools contain 15-mer peptides corresponding to immunodominant epitopes. Peptides stimulate both CD4 and CD8 T cells. Each 60nmoL vial was reconstituted aseptically in 2mL of sterile de-ionized water and all 4 vials were combined into a single mega pool. For in-tube stimulation 80µL of the mega pool was dispensed into a BD vacutainer no additive tube (Beckton Dickenson; city, state). Phytohemagglutinin PHA-P Mitogen (Sigma) was dissolved in sterile deionized water at 1mg/mL. For in-tube stimulation, 50µL was dispensed into a BD Vacutainer No Additive Tube. Prepared tubes and peptide reagents were stored at -80 °C.

### Interferon Gamma Release Assay (IGRA)

SARS-CoV-2 IGRA was performed as described previously [12]. One mL of freshly collected blood was transferred to a 24 well tissue culture plate or BD vacutainer with no additive tubes at 1mL per well/tube. One well or tube was left unstimulated (nil), one well or tube was stimulated with SARS-CoV-2 antigen mega pool at 2.2 mmol/mL, and one well or tube was stimulated with mitogen at 50µL/mL. In-tube stimulation was validated using in-plate stimulation as the reference method (Supplementary Figure 1). The blood samples were mixed gently and incubated at 37 °C with a relative humidity of 95% for 20 to 24 h. Plasma was separated and stored at 4 °C. The IFN-γ concentration was measured with an automated enzyme-linked immunosorbent assay (ELISA) instrument (DSX; Dynex Technologies, Chantilly, VA) using the QuantiFERON-TB ELISA kit (Qiagen, Germantown, MD)). A four-point standard curve was used to calculate IFN-γ concentration in international units (IU)/mL. IFN-γ response was defined as positive if antigen-nil ≥0.35 IU/mL; negative if antigen-nil <0.35 and mitogen-nil ≥0.5 IU/mL, and indeterminate if nil >8 IU/mL or antigen-nil <0.35 and mitogen-nil <0.5 IU/mL.

### Enzyme-linked immunosorbent assay (ELISA)

Anti-SARS-CoV-2 Spike S1 domain IgG ELISA was performed on lithium heparin plasma using the EUROIMMUN instrument and reagents (Lübeck, Germany) per the manufacturer’s instructions. The anti-SARS-CoV-2 IgG antibody level was reported as the ratio of optical density (OD) of the sample over the OD of the calibrator. The ratio was defined as follows: <0.8 negative; ≥0.8 to <1.1 borderline; ≥1.1 positive.

### Statistical Analysis

The Mann-Whitney U test was used to compare median IFN-γ response between groups. Fisher’s exact test was used to analyze differences between proportions. Statistical analysis was done with GraphPad Prism 8.0.1 software (San Diego, CA, USA).

## Results

### Patient Cohorts

To assess the accuracy of SARS-CoV-2 IGRA longitudinally after natural infection, a total of 94 unique adult COVID-19 convalescents were tested at 0.5 (n=16), 1 (n=8), 4 (n=48), 7 (n=70) and 10 (n=39) months post infection for total of 181 tests. A total of 142 uninfected healthy blood donors were tested one time. COVID-19 convalescents had either mild (91%) or asymptomatic (9%) COVID-19. The median age of COVID-19 convalescents was 38 years (interquartile range IQR], 29-55 years) and 45% were female. To evaluate the utility of SARS-CoV-2 IGRA in household contacts on index cases with COVID-19, 12 index cases (index cases from 2 households did not participate) and 23 contacts belonging to 14 households were tested. Demographic and clinical characteristics of the household participants are summarized in Table 1 and Supplementary Table 1. The median age of household participants was 46 years (IQR, 20-62 years) and 49% were female. The median time between index case RT-PCR positivity and blood collection for this study was 17 days (IQR, 5-96 days). Six contacts (26.1%) were RT-PCR positive after exposure to index case but; four of them were RT-PCR positive on the day of sample collection for this study.

**Table 1.** Demographic and clinical characteristics of house hold members.

| Demographic and Clinical Characteristics |  | Index cases (n=12) |  | Contacts (n=23) |  |
| --- | --- | --- | --- | --- | --- |
|  |  | n | % | n | % |
| Median age (IQR) |  | 56 (24-66) |  | 44 (16-54) |  |
| Male |  | 7 | 58.3 | 11 | 47.8 |
| Race | White | 8 | 67.7 | 17 | 73.9 |
|  | Hispanic | 4 | 33.3 | 4 | 17.4 |
|  | Asian | - | - | 2 | 8.7 |
| Symptoms | Fever | 4 | 33.3 | 2 | 8.7 |
|  | Cough | 6 | 50.0 | 6 | 26.1 |
|  | Sore throat | 3 | 25.0 | 3 | 13.0 |
|  | Running nose | 6 | 50.0 | 3 | 13.0 |
|  | Chest pain | 3 | 25.0 | 1 | 4.3 |
|  | Joint pain | 1 | 8.3 | 2 | 8.7 |
|  | Fatigue | 4 | 33.3 | 5 | 21.7 |
|  | Shortness of breath | 3 | 25.0 | 2 | 8.7 |
|  | Loss of smell | 2 | 17.7 | 1 | 4.3 |
|  | Diarrhea | - | - | 1 | 4.3 |
|  | Headache | 1 | 8.3 | 3 | 13.0 |

### Accuracy of SARS-CoV-2 IGRA

In COVID-19 convalescents tested with SARS-CoV-2 IGRA, the overall sensitivity over 10 months post-infection was 84.5% (153/181; 95% confidence interval [CI] 79.0%–89.0%) (Table 2 and Figure 2A). When only their first test result is considered, the overall sensitivity was 85.1% (80/94; 95% CI 76.4–91.0). In healthy blood donors the specificity was 86.6% (123/142; 95% CI 80.0-91.2) (Table 2 and Figure 2A). No indeterminate results were obtained in both groups. The sensitivity at 0.5 and 1-month post infection was 100% and dropped to 79.5% (P<0.01) at 10-months post infection (Table 2 and Figure 2A). Compared with median IFN-γ response in healthy blood donors (0.02 IU/mL, IQR 0.01-0.06), the IFN-γ response was significantly higher in convalescents at 0.5-month (3.77 IU/mL, IQR 1.38-6.75, P<0.001), 1-month (2.96 IU/mL, IQR 1.31-6.10, P<0.001), 4-month (2.15 IU/mL, IQR 0.54-3.53, P<0.001), 7-month (1.16 IU/mL, IQR 0.53-3.86, P<0.001), and 10-month (1.26 IU/mL, IQR 0.38-2.96, P<0.001) post-infection (Figure 2A). Among convalescents, compared with 0.5-month post-infection, IFN-γ response was not significantly different at 1 and 4-month post-infection (Figure 2A). However, there was a significant decline in IFN-γ response at 7-month (P<0.05), and 10-month (P<0.01) post-infection (Figure 2A).

**Table 2.** Accuracy of SARS-CoV-2 IGRA and IgG in convalescents and healthy blood donors.

|  |  | Months Since SARS-CoV-2 Infection (No. of Subjects) |  |  |  |  | Health Blood Donors<br>(No. of Subjects) |
| --- | --- | --- | --- | --- | --- | --- | --- |
|  |  | 0.5 (16) | 1 (8) | 4 (48) | 7 (70) | 10 (39) | 142 |
| IGRA | **Sensitivity** | 100 (16/16; | 100 (8/8; | 85.4 (41/48; | 81.4 (57/70; | 79.5 (31/39; | - |
|  | **(n/N; 95% CI)** | 80.6-100) | 67.6-100) | 72.8-92.8) | 70.7-88.8) | 64.5-89.2) | - |
|  | **Specificity** | - | - | - | - | - | 86.6 (123/142; |
|  | **(n/N; 95% CI)** | - | - | - | - | - | 80.0-91.2) |
| IgG | **Sensitivity** | 87.5 (14/16; | 100 (8/8; | 87.5 (42/48; | 85.7 (60/70; | 71.8 (28/39; | - |
|  | **(n/N; 95% CI)** | 63.9-97.8) | 67.6-100) | 75.3-94.1) | 75.7-92.1) | 56.2-83.5) | - |
|  | **Specificity** | - | - | - | - | - | 97.8 (3/181; |
|  | **(n/N; 95% CI)** | - | - | - | - | - | 93.9-99.4) |

No, number; CI, confidence interval

**Figure 2.**
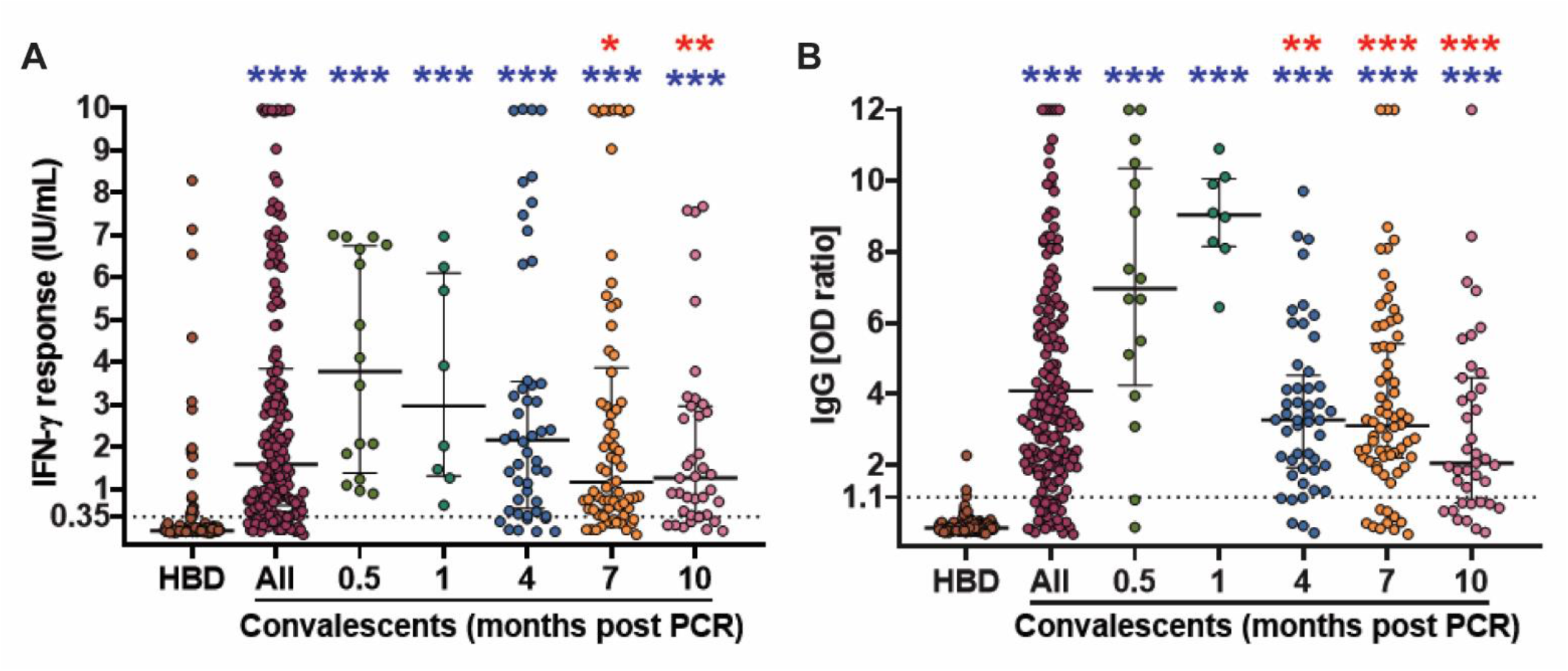
SARS-CoV-2 IGRA and IgG ELISA results in convalescents and healthy blood donors. IFN-γ response with whole blood IGRA (A) and IgG antibody optical density (OD) ratio (B) in healthy blood donors (HBD) (n=142) and COVID-19 convalescents at 0.5 (n=16), 1 (n=8), 4 (n=48), 7 (n=70), and 10 (n=39) months post-infection. Dotted lines represent the assay cutoffs (0.35 IU/mL for IGRA and 1.1 OD ratio for IgG). Quantitative results >10 IU/mL are shown as 10 IU/mL and >12 OD ratio are shown as 12 OD ratio. Horizontal lines show the median. whiskers show the range of IFN-γ response (IU/mL). Response in convalescents was compared to controls (blue). Response in convalescents were compared to 0.5 month time point (red) *, P < 0.05; **, P < 0.01; ***, P <0.001.

The overall sensitivity of IgG over 10 months post-infection was 83.9% (152/181; 95% CI 77.9-88.6) in convalescents. The specificity was 97.8% (3/181; 95% CI 93.9-99.4) in healthy blood donors (Table 2 and Figure 2B). The sensitivity was high at 0.5-month (87.5% [14/16]; 95% CI 63.9-97.8) and 1-month (100% [8/8]; 95% CI 67.6-100) post infection compared with 71.8% (28/39; 95% CI 56.2-83.5; P<0.001) at 10-month (Figure 2B). Compared with median IgG OD ratio in healthy blood donors 0.24 (IQR 0.18-0.32) the median IgG OD ratio was 6.97 (IQR 4.25-10.36, P<0.001) at 0.5-month, 9.04 (IQR 8.14-10.06, P<0.001) at 1-month, 3.27 (IQR 1.94-4.52, P<0.001) at 4-month, 3.11 (IQR 2.2-5.42, P<0.001) at 7-month, and 2.07 (IQR 0.94-4.46, P<0.001) at 10-month time points (Figure 2B). Compared with 0.5-month post-infection, IgG OD ratio was significantly lower at 4 (P<0.01), 7 (P<0.001), and 10-month (P<0.001) post-infection (Figure 2B).

When comparing IGRA with IgG, the overall concordance between qualitative results was 79.6% (144/181) in convalescents and 85.9% (122/142) in healthy blood donors. Quantitatively, no significant correlation was found between IGRA IFN-γ response and IgG OD ratio (r=0.11, P>0.05) (Supplementary Figure 2).

### SARS-CoV-2 IGRA in Household contacts

In household members tested one time after recruitment, 100% (12/12) of the index cases and 65.2% (15/23) of the contacts were positive with the SARS-CoV-2 IGRA (Figure 3A). No indeterminate results were obtained. In 13 contacts with history of RT-PCR-positive SARS-CoV-2 infection (7 with remote history), 12 (92.3%) were SARS-CoV-2 IGRA positive. The median IFN-γ response was 2.81 IU/mL (IQR, 0.67-5.73) in the index cases compared with 1.38 IU/mL (IQR, 0.03-2.7, P≥0.05) in the contacts (Supplementary Figure 3A).

**Figure 3.**
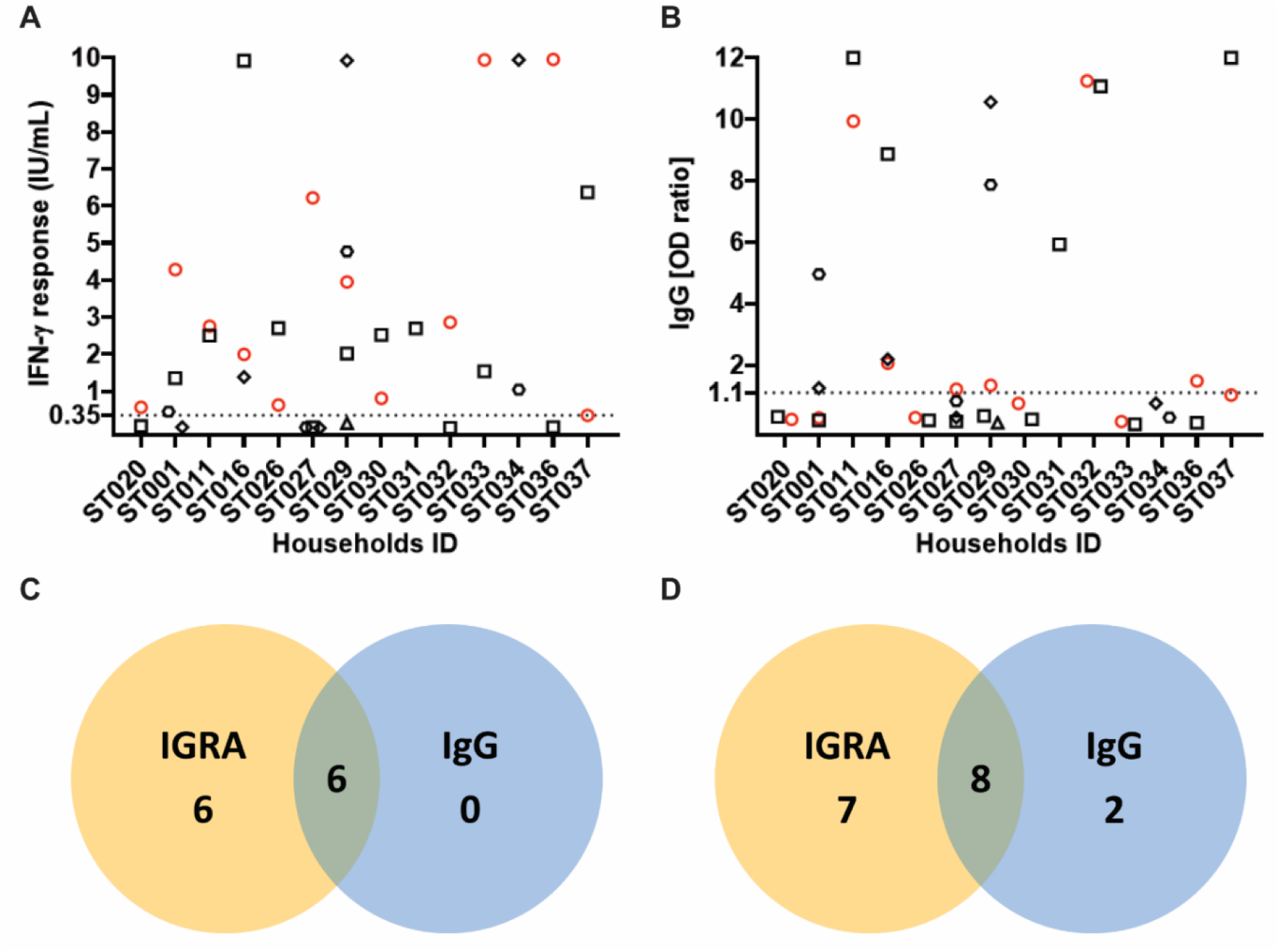
SARS-CoV-2 IGRA and IgG ELISA result in index cases and household contacts. IFN-γ response with whole blood IGRA (A) and IgG antibody optical density (OD) ratio (B) in 14 households with 12 index cases (red circles) and 23 contacts (open black symbols). Venn diagrams show positive results with IGRA and/or IgG in index cases (C) and contacts (D). Dotted lines represent the assay cutoffs (0.35 IU/mL for IGRA and 1.1 OD ratio for IgG). Quantitative results >10 IU/mL are shown as 10 IU/mL and >12 OD ratio are shown as 12 OD ratio.

SARS-CoV-2 IgG ELISA was positive in 50.0% (6/12) of the index cases and 43.5% (10/23) of the contacts (Figure 3B). In 13 contacts with history of SARS-CoV-2 infection, 6 (46.2%) were SARS-CoV-2 IGRA positive. The median IgG antibody ratio was 1.11 (IQR, 0.28-1.91) in the index cases compared with 0.75 (IQR, 0.2-7.87, P>0.05) in the contacts (Supplementary Figure 3B).

In the household cohort, concordance between IGRA and IgG results was 50.0% (6/12) and 60.9% (14/23) in the index cases and the contacts, respectively. When combining IGRA and IgG results, either test was positive in 100% (12/12) of the index cases and 73.9% (17/23) of the contacts (Figure 3C and 3D). Compared with IgG, IGRA had a differential positivity rate of +50% (95% CI +25.4-+74.6) and +21.7% (95% CI, +9.23-+42.3) in the index cases and the contacts, respectively. The positivity rate of IGRA was significantly higher than IgG in index cases and contacts (P<0.05 for both).

## Discussion

Accurate immunodiagnostics are needed to assess T cell-mediated immune response to SARS-CoV-2 after natural infection and after vaccination to inform providers on possible protective immunity to SARS-CoV-2 [1] and to accurately understand transmission dynamics of emerging variants. Furthermore, T cell responses may be important for understanding immunity to SARS-CoV-2 infection, particularly in the context of emerging variants that might escape antibody responses[15, 16]. Using a simple in-tube whole blood SARS-CoV-2 IGRA, we show that T cell-mediated immune response to SARS-CoV-2 was sustained longitudinally with 85% and 80% sensitivity at 4- and 10-month, respectively, post-infection in convalescents who had mild or asymptomatic COVID-19. Our findings are consistent with a prior study showing a robust memory T-cell response months after SARS-CoV-2 infection in individuals with mild or asymptomatic infection [17, 18], and similar to prior studies we observed a mild decline in the cellular and humoral immune response in convalescents [19-21]. Consistent with prior studies we also found a higher proportion of household contacts with positive SARS-CoV-2 IGRA compared with IgG [6, 7]. These findings indicate that the SARS-CoV-2 IGRA used in this study is more sensitive than IgG testing for detection of asymptomatic or mild infection amongst close contacts, at least in the early period, and thus may be an important tool for epidemiological studies aimed at acutely understanding the transmission dynamics of emerging SARS-CoV-2 variant such as the delta variant [22].

SARS-CoV-2 IGRA may also serve an important role in the clinical assessment of T-cell-mediated immune response to the SARS-CoV-2 vaccines. Humoral immune response has been shown to decline after SARS-CoV-2 vaccination [19-21]. Furthermore, it was recently shown that the antibody response rate to the SARS-CoV-2 vaccine in transplant patients was low after the second dose (40%) and third dose (68%)[23]. Preliminary analysis of SARS-CoV-2 vaccine response in immunocompromised patients at our health system has revealed a significantly higher IGRA positivity rate compared with IgG positivity rate (unpublished data). These findings support the role of SARS-CoV-2 IGRA in vaccine response assessment, particularly in elderly and immunocompromised patients given that immunosenescence and immunosuppression, respectively, may dampen adaptive immune responses and leave the host vulnerable to subsequent infection [3, 24, 25]. Thus, IGRA and IgG serology may serve a role in informing providers on the status of cell-mediated and humoral response to SARS-CoV-2 vaccine and the need for revaccination.

Although the findings are promising, this study has several limitations. First, the whole blood SARS-CoV-2 IGRA did not distinguish between CD4 and CD8 T cell responses. Prior studies in convalescents have shown that IFN-γ response in IGRA is predominantly CD4 T cell-derived [12, 26]. However, a more complex assay design that allows measurement of CD4 and CD8 T cells response is possible if clinically indicated. Second, given that healthy blood donors were recruited during the pandemic, we would not know whether the 16% positivity rate with SARS-CoV-2 IGRA in this group was due to true-positive results due to past SARS-CoV-2 infection, false-positive results, or cross-reactivity due to past infection with seasonal CoV [7, 27, 28]. Third, the IgG response in this study was not further assessed for its ability to neutralize the virus. Such characterization was not relevant in the context of investigating IGRA accuracy. Forth, SARS-CoV-2 IGRA was not applied to the investigation of vaccine response in this study. Studies are underway to measure the vaccine response with IGRA in immunocompromised patients. Lastly, the household study lacked multiple time points to more accurately assess the performance of IGRA vs IgG, and we did not perform risk assessment in household contacts to correlate IGRA positivity with exposure risk.

In summary, the whole blood SARS-CoV-2 IGRA was shown to maintain sensitivity in convalescents up to 10 months post-infection and was shown to have a higher positivity rate than IgG in household contacts of COVID-19 cases. SARS-CoV-2 IGRA is a simple and robust clinical immunodiagnostic test that can be applied to accurately understand the transmission of emerging variants and to assess vaccine response in vulnerable populations.

## Data Availability

The data used to support the findings of this study are included within the article.

